# Moving psychiatric deinstitutionalisation forward: A scoping review of barriers and facilitators

**DOI:** 10.1101/2023.03.28.23287810

**Authors:** Cristian Montenegro Cortés, Josefa González Moller, Matías Irarrázaval Dominguez, Felicity Thomas, Jorge Urrutia Ortiz

## Abstract

Psychiatric deinstitutionalisation (PDI) processes aim to transform long-term psychiatric care by closing or reducing psychiatric hospitals, reallocating beds, and establishing comprehensive community-based services for individuals with severe and persistent mental health difficulties. This scoping review explores the extensive literature on PDI, spanning decades, regions, socio-political contexts, and disciplines, to identify barriers and facilitators of PDI implementation, providing researchers and policymakers with a categorization of these factors.

To identify barriers and facilitators, three electronic databases (Medline, CINAHL, and Sociological Abstracts) were searched, yielding 2250 references. After screening and reviewing, 52 studies were included in the final analysis. Thematic synthesis was utilized to categorize the identified factors, responding to the review question.

The analysis revealed that barriers to PDI include inadequate planning, funding, and leadership, limited knowledge, competing interests, insufficient community-based alternatives, and resistance from the workforce, community, and family/caregivers. In contrast, facilitators encompass careful planning, financing and coordination, available research and evidence, strong and sustained advocacy, comprehensive community services, and a well-trained workforce engaged in the process. Exogenous factors, such as conflict and humanitarian disasters, can also play a role in PDI processes.

Implementing PDI requires a multifaceted strategy, strong leadership, diverse stakeholder participation, and long-term political and financial support. Understanding local needs and forces is crucial, and studying PDI necessitates methodological flexibility and sensitivity to contextual variation. At the same time, based on the development of the review itself, we identify four limitations in the literature, concerning ‘time’, ‘location’, ‘focus’, and ‘voice’. We call for a renewed research and advocacy agenda around this neglected aspect of contemporary global mental health policy is needed.

**Impact Statement:** The transition from a mental health system centred on long-term psychiatric hospital care to one centred on community-based services is complex, usually prolonged and requires adequate planning, sustained support and careful intersectoral coordination. The literature documenting and discussing psychiatric Deinstitutionalisation (PDI) processes is vast, running across different time periods, regions, socio-political circumstances, and disciplines, and involving diverse models of institutionalisation and community-based care. This scoping review maps this literature, identifying barriers and facilitators for PDI processes, developing a categorization that can help researchers and policymakers approach the various sources of complexity involved in this policy process.

Based on the review, we propose five key areas of consideration for policymakers involved in PDI efforts: 1) Needs assessment, design and scaling up; 2) Financing the transition. 3) Workforce attitudes and development; 4) PDI Implementation and 5) Monitoring and quality assurance.

We call for a multifaceted transition strategy that includes clear and strong leadership, participation from diverse stakeholders and long-term political and financial commitment. Countries going through the transition and those who are starting the process need a detailed understanding of their specific needs and contextual features at the legal, institutional, and political levels.

## Introduction

Starting during and after World War II in Western Europe and North America, psychiatric deinstitutionalisation (PDI) is widely considered a central element of the modernization of psychiatry. It involves two broad components: (i) the closure or reduction of large psychiatric hospitals and (ii) the development of comprehensive community-based mental health services aiming to promote social inclusion and full citizenship for people living with severe mental illness A broad international consensus supports the need for a shift in mental health care, away from long-term institutionalisation and towards comprehensive and integrated community-based and community-shaped services (Campbell & Burgess, 2012; Thornicroft et al., 2016; WHO, 2013, 2021a)

Significant economic, social, and cultural forces have precipitated the development of PDI, including public awareness of the dehumanizing effects of prolonged institutionalisation in often poor conditions, the high cost of maintaining large, long-stay institutions, and pharmaceutical developments such as the introduction of psychotropic medication (Salisbury et al., 2016; Turner, 2004; Yohanna, 2013). For several decades, advocacy movements across the mental health and disability fields have demanded the protection of patients’ human rights, including the right to live independently in the community (Hillman, 2005; Mezzina et al., 2019). The UK, Italy, and Finland among other countries are generally regarded as good examples of PDI (Barbui et al., 2018; Turner, 2004; Westman et al., 2012). In the global south, while varying in approach and scale, Brazil, Chile, Sri Lanka and Vietnam have received praise for their efforts to move away from centralized psychiatric institutions (PAHO, 2008; Cohen & Minas 2017).

Despite the consensus and the declarations by many governments, PDI remains a complex, and fragile endeavour. Progress towards PDI varies greatly across and within countries (Hudson, 2019). In some regions, the majority of resources are still invested on centralized, long term hospitalization (WHO and the Gulbenkian GMHP, 2014); in others, PDI has been delayed with the balance of mental health care shifting in favour of hospital-focused care (Sade et al., 2021); and in other cases, poor management of the PDI process has resulted in tragedy (see for example Moseneke’s 2018 account of the Esidimeni tragedy in South Africa).

Understanding the factors that lead to or prevent the transition is crucial to inform the planning and implementation of PDI. Whilst these factors have been documented through the accounts of leaders and experts with hands-on experience, such as in the WHO’s Innovation in Deinstitutionalisation report (WHO and the Gulbenkian GMHP, 2014), there has been no previous attempt to systematically scope the literature on barriers and facilitators to PDI.

This paper therefore reports the results of a Scoping Review examining the extent and range of available research regarding barriers and facilitators involved in PDI processes. We organised the specific barriers in seven groups, and the facilitators in six groups, totalling thirteen thematic groups. This categorization can be adapted to national realities and different levels of policy action around PDI, to guide research and policy efforts. The synthesis of this information allows us to establish a list of suggestions on ways to move forward.

## Methods

Given that the literature on this topic has not been comprehensively reviewed, the Scoping Review (ScR) (Arksey & O’Malley, 2005) methodology was used. The goal of a ScR is “to map rapidly the key concepts underpinning a research area and the main sources and types of evidence available (…), especially where an area is complex or has not been reviewed comprehensively before” (Mays et al., 2001, p. 194). For this review, a barrier to PDI was defined as any factor limiting or restricting the transition of care from long-term hospitalization to community-based services and supports. This may include, but is not limited to, issues related to the public-health priority agenda (Shen & Snowden, 2014); challenges in the implementation of mental health services in community settings (Kormann & Petronko, 2004; Saraceno et al., 2007); the resistance of workers employed by psychiatric institutions (Priebe 2002); and public and community responses, including stigma, paternalism and other sociocultural factors (Fisher et al., 2005; O’Doherty et al., 2016).

Correspondingly, we define a facilitator as any factor that fosters, promotes, or enables an adequate PDI process. These include the presence of well-organised social activism supporting the rights of persons with mental health problems (Anderson et al., 1998), the acceptance of mental illness as a human condition (Gostin, 2008), service paradigms that enhance social inclusion and citizenship (Fakhoury & Priebe, 2002; Saraceno, 2003) and political willingness (Saraceno et al., 2007).

This ScR was conducted following the Checklist for Preferred Reporting Items for Systematic reviews and Meta-Analyses extension for Scoping Review (PRISMA-ScR) (Tricco et al., 2018). A review protocol was created and registered at the Open Science Platform (doi: 10.17605/OSF.IO/XEBQW). See the protocol and PRISMA-ScR Checklist in Supplementary materials A and B, respectively.

Three electronic databases were searched in May 2020 - Medline, CINAHL and Sociological Abstracts. Previously published systematic reviews on adults with severe mental health impairment (Lean et al., 2019; Richardson et al., 2019), barriers and facilitators to healthcare access (Adauy et al., 2013) and the Deinstitutionalisation process (May et al., 2019) informed our search strategy. The strategy combined terms across three dimensions: (i) adults with mental health impairment; (ii) barriers and facilitators related to health care delivery; and (iii) the Deinstitutionalisation process. The search strategy was not limited by study design or country. Tailored searches were developed for each database (see Supplementary material C). Eligibility criteria was limited by studies in English and Spanish. All references obtained through the electronic database search and hand search were pooled in EndNote 11 (reference manager) and then uploaded to Covidence (screening and data extraction tool).

Studies selected for inclusion met the criteria detailed in Table 1. Initial eligibility was independently assessed by JU and JG based on title and abstract. At the level of full-text screening, a random sampling of 10% of the selected studies was pilot-tested (with three reviewers) to ensure at least 80% of agreement. Differences in opinions were discussed, and a final decision on their eligibility was made after discussion with CM. A specific data extraction form was created to record full study details and guide decisions about the relevance of individual studies (Table 2). Two reviewers (JU and JG) extracted data and checked for accuracy with another reviewer (CM). Eligibility criteria was further specified to differentiate and exclude specialized substance abuse services involving the legal system. Studies on child institutionalisation and substance abuse were also excluded because of the distinct set of causes and challenges associated with these phenomena. Articles related to transinstitutionalisation, the transfer of users from psychiatric hospitals to other institutional settings were excluded unless they addressed PDI barriers and facilitators directly.

During the research process, inclusion criteria adopted a dimensional character, with studies clearly stating barriers and facilitators on one extreme and studies where they had to be inferred, on the other. Given that ScR methodology is defined as an exploratory strategy to map the state of research on a topic (Arksey & O’Malley, 2005; Peters et al., 2015), no attempts were made to assess the methodological quality of the included studies.

Thematic synthesis (Harden, 2010; Lucas et al., 2007; Thomas et al., 2004; Thomas & Harden, 2008) of the selected papers followed a three-stage process. Firstly, it involved free coding the content of the text, to identify barriers and facilitators. Secondly, grouping and organizing the codes into an inductively developed set of categories. Finally, CM examined the categories and their respective codes in the light of the review question to produce an initial set of categories. The match between codes (barriers/facilitators) and categories, and their relevance for the review question was further discussed and refined through rounds of collective revision. A table with examples of the data coding process is available (Supplementary Material D).

To consistently scope the academic production around PDI over several decades, this review includes publications up until May 2020, intentionally excluding the literature related to the Covid-19 pandemic. To properly assess the effects of the Covid-19 pandemic upon processes of Deinstitutionalisation -and on the reality of long-term psychiatric hospitals in general- a different research question, and a tailored design is required.

## Results

The search strategy retrieved 2250 references. Nine more references were added after hand-searching reference lists and contacting relevant authors. After duplicate removal, 1915 references were screened by title and abstract, leaving 215 articles for full-text screening. Finally, 52 studies were included in the analysis. Search results and the reasons for excluding full-text articles are provided in the PRISMA flowchart (Figure 1).

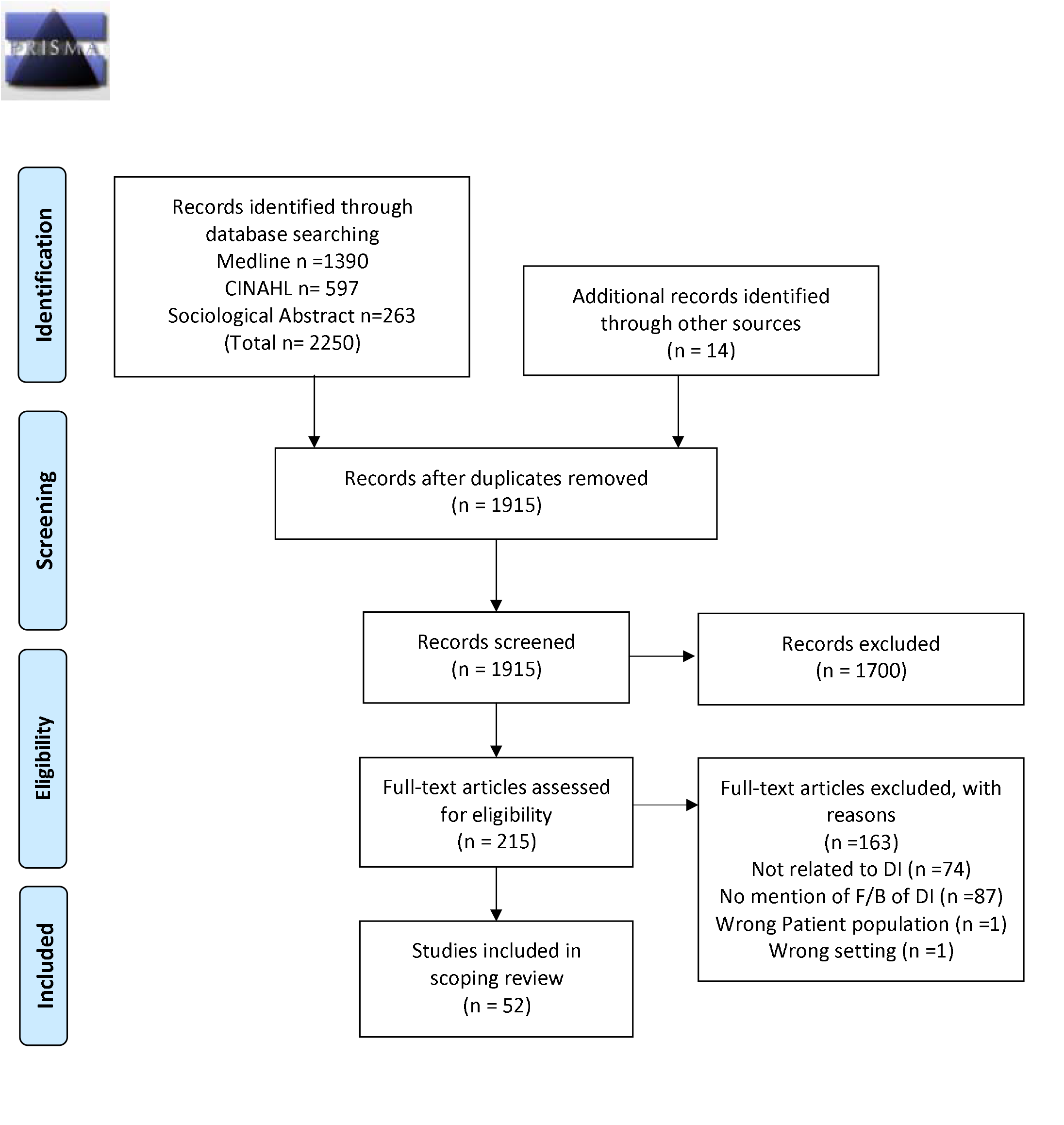
PRISMA 2009 Flow Diagram. *From:* Moher D, Liberati A, Tetzlaff J, Altman DG, The PRISMA Group (2009). *P*referred *R*eporting *I*tems for *S*ystematic Reviews and *M*eta-*A*nalyses: The PRISMA Statement. PLoS Med 6(7): e1000097. doi:10.1371/journal.pmed1000097 For more information, visit www.prisma-statement.org.

### Characteristics of the studies

Included studies were published between 1977 and 2019. This broad temporal scope responds to the fact that an important proportion of research was parallel to the implementation of PDI policies in Europe and the USA during the 1970s and 1980s. Studies were predominantly conducted in the USA (n=22), followed by the UK (n=7) and Canada (n=5). Figure 2 shows an overview of the geographical distribution of the included studies. Regarding the methodology, 25 publications were qualitative studies, 22 were quantitative, and 5 used mixed methods. We provide a summary of the studies’ characteristics in Table 3 and descriptions of each study in Table 4.

**Figure 2.**
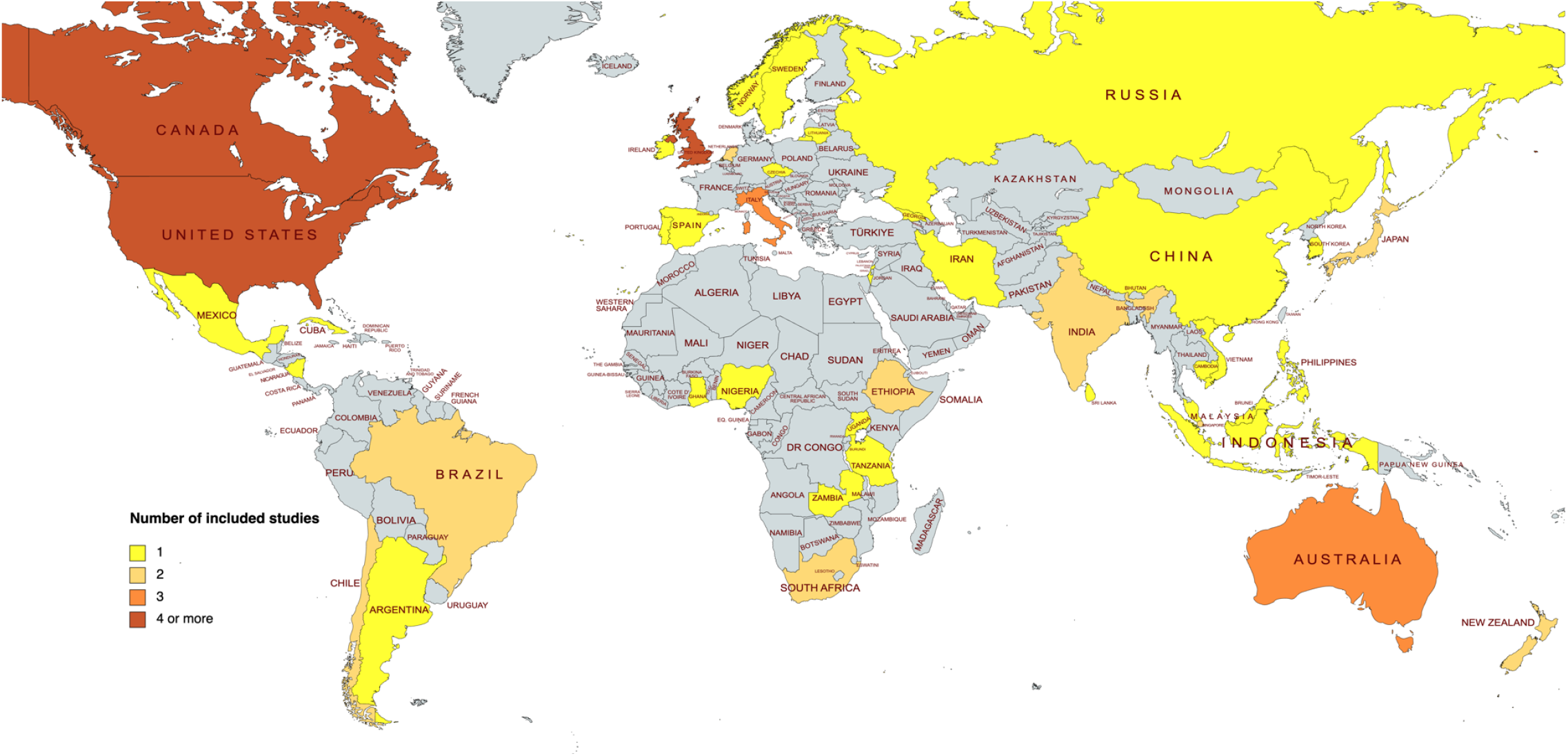
Geographical distribution of included studies *The following countries were included in one or more multi-country studies: Malaysia, Japan, Ethiopia, Brazil, Nigeria, Uganda, UK, Iran, Italy, Portugal, Cambodia, Philippines, Spain, New Zealand, Usa, Sri Lanka, Chile, India, Republic of Korea, The Netherlandands, Zambia, Indonesia, Tanzania, Singapore, Lithuania, Australia, Georgia, Vietnam, South Africa, Ghana, Sweden, Argentina, Cuba, Jamaica and Mexico.

It is important to consider that this is a general categorization based on the available literature, whose aim is to identify what has been reported as a barrier and as a facilitator in a systematically selected, diverse set of references. We applied thematic analysis to the entire set, and on that basis, we developed this initial categorization. We are not establishing the prevalence of each barrier/facilitator across the set or contrasting the characteristics of each barrier/facilitator across regions or within a specific stage in the PDI process. For specific information about the composition of the categories and codes, see table 5 for barriers and table 6 for facilitators.

### Barriers to the process of psychiatric Deinstitutionalisation

Barriers to the process were organised under seven categories, summarized in Table 5 and described in detail below.

#### 1. Planning, leadership, and funding

This category includes barriers related to design, implementation, monitoring and overall leadership of the process, and its interaction with other policy processes. One barrier is the lack of accountability from the government to carry out the reform properly, refusing responsibility for housing, social or medical needs and not including other agencies in patient discharge planning (Rose, 1979). The absence of clear operational goals may hinder performance evaluation of the process (Rosenheck, 2000). Charismatic and ideologically driven leadership of the process is important at the beginning, although is vulnerable to political shifts, including elections and changes in government (PAHO, 2008).

Barriers related to funding included the lack of a clear policy that assured the reallocation of resources from hospitals to CMHS (Fakhoury & Priebe, 2002; PAHO, 2008) and a lack of funding to ensure the continuity of community services (McCubbin, 1994; Mechanic & Rochefort, 1990; PAHO, 2008). This to secure a synchronicity between downsizing psychiatric hospitals and the scaling up of psychosocial interventions.

#### 2. Knowledge/Science

Conceptual barriers to promoting PDI were identified. Some authors consider that the lack of research on PDI processes (Bennett & Morris, 1981), paralyze or slow down policy planning and implementation (Shen & Snowden, 2014). At the conceptual level, reducing the concept of community care to narrow geographical proximity can limit the development of community-based interventions (Bennett & Morris, 1983).

Some authors criticised the inadequate transfer and use of certain service paradigms, such as the application of urban-centred interventions to rural locations (Kraudy et al., 1983) without previous identification of rural specificities, creating a disconnection between users and facilities (Schmidt, 2000).

#### 3. Power, interests, and influences

Barriers related to the conflict between the interests and perspectives of different groups were grouped under this category.

Authors have discussed the impact of the privatization of mental health care in the wake of the closure of psychiatric hospitals. Market-driven decisions can re-create similar conditions to those in old psychiatric facilities (Rose, 1979). The rise of private hospitals in the United States and their reluctance to participate in non-profit services, such as working with existing public providers, influences access to and the nature of mental health care. Private for-profit hospitals may restrict access to care for uninsured patients (Dorwart et al., 1991). Additionally, private insurance in the United States often encourages unnecessary hospitalization and discourages psychosocial interventions and alternative forms of treatment (Freedman & Moran, 1984; Barton, 1983).

Furthermore, the low cost of hospitalization in some areas, as reported in Asia (Fakhoury & Priebe, 2002), does not provide an economic incentive to push for deinstitutionalization”.

The dependence of psychiatric research and development on drug-companies is seen as a barrier. McCubbin stated that the vested interests of the pharmaceutical industry may influence psychiatric practice by selectively supporting medical schools, conferences, and journals, potentially tuning the vision of community mental health into a market opportunity (McCubbin, 1994).

Finally, the lack of relevance of mental health in the political agenda is a crucial, over-encompassing barrier to effective advocacy efforts (Mechanic & Rochefort, 1990; PAHO, 2008; Semke, 1999), as is the uncoordinated and fragmentary nature of these efforts (McCubbin, 1994; Mechanic & Rochefort, 1990; Rosenheck, 2000).

#### 4. Services and support in the community

The slow development of community programmes forced patients to return to long-term institutions, risking chronification (Kaffman et al., 1996). There have been reports of problems caused by the sudden decrease in psychiatric beds without corresponding increases in community-based services. This can result in unintended transfers of patients to other institution-based services and even imprisonment (Shen & Snowden, 2014). Inadequate training of community-based workers, discharge without community support (Shen & Snowden, 2014) and early release promoted by legislatively mandated PDI policies (Kleiner & Drews, 1992) are elements to consider.

The authors identified several barriers to adequate integration of discharged users into their communities, including the absence of jobs and income (Goering, 1984), inadequate housing (Grabowski et al., 2009), and insufficient public support (Manuel et al., 2012). Other barriers included challenging behaviors (Allen et al., 2007), old age (Barry et al., 2002), and pessimistic attitudes and feelings of disempowerment and hopelessness among patients (Chopra & Herrman, 2011). In addition, the decrease in disability pensions following an increase in earned income was also identified as a barrier to social integration, as it can discourage work (Chopra & Herrman, 2011).”

#### 5. Workforce

Barriers related to the workforce in both institutionalized settings and community services were identified. Regarding human resources, authors mentioned staff shortages as a barrier for the transition towards community-based care (Fakhoury & Priebe, 2002; Rose, 1979; Shen & Snowden, 2014, Stelovich, 1979). Another barrier reported was the internal frictions and the existence of opposing views about care and rehabilitation (Kaffman et al., 1996; O’Doherty et al., 2016). More specifically, the psychiatric hospital workforce can delay or hinder the transformation of psychiatric institutions for fear of losing their livelihoods (Shen & Snowden, 2014; Swidler & Tauriello, 1995). Workers can express reluctance and scepticism regarding the feasibility of community living for institutionalized persons (Mayston et al., 2016; O’Doherty et al., 2016). This includes the development of unfair expectations toward family members, which alienated carers and hindered their willingness to accept responsibility (Barton, 1983).

On the other hand, service providers located in the community can be sources of stigma, expressed in the avoidance of formerly institutionalized patients (Barton, 1983), hopelessness towards treatment (Aggett & Goldberg, 2005), exclusion of users from constructing their treatment plan (Bryant et al., 2004) and fears stemming from the lack of restraining measures (Ash et al., 2015). Perceived racism at the hands of service providers can lead to mistrust in patients, causing them to either reject treatment or have poor adherence, which in turn can result in poorer outcomes, such as a longer hospital stays (Chakraborty et al., 2011).

#### 6. Communities and the public

Factors limiting social inclusion, comprising attitudes towards persons with SMI and community responses to PDI processes, were grouped under this category. Lack of preparation and stigma (Aggett & Goldberg, 2005; Bredenberg, 1983; Chan & Mak, 2014; Fakhoury & Priebe, 2002; Manuel et al., 2012; Mechanic & Rochefort, 1990; O’Doherty et al., 2016; PAHO, 2008) leads to hostile attitudes toward service-users challenging social integration (Aggett & Goldberg, 2005; Bredenberg, 1983; Fakhoury & Priebe, 2002; O’Doherty et al., 2016, PAHO, 2008). The attribution of dangerousness to individuals with SMI and the public acceptance of social control measures over recovery-oriented alternatives were also reported as barriers to PDI processes (Fakhoury & Priebe, 2002; Matsea et al., 2019)

#### 7. Family/Carers

Authors highlighted the difficulties in maintaining relationships between caregivers and community services (Aggett & Goldberg, 2005; Barton, 1983; Yip, 2006; Lavoie-Tremblay et al., 2012; McCubbin, 1994; Mayston et al., 2016; O’Doherty et al., 2016). Previous experiences of failed treatments can lead to lack of cooperation and hostility towards services (Aggett & Goldberg, 2005). Professionals can be reluctant to cooperate and skeptical about the feasibility of community living (Mayston et al., 2016; O’Doherty et al., 2016). Families and caregivers may have concerns about community living and its suitability for people with high support needs (O’Doherty et al., 2016) and concerns about receiving the burden of care, and this can alienate them and hinder their willingness to accept responsibility.

### Facilitators to the process of psychiatric Deinstitutionalisation

Facilitators in the process were organised under six categories summarized in Table 6 and described in detail below.

#### 1. Planning, leadership, and funding

Factors related to organizational and managerial capacities required for the transition were grouped under this category. Authors stated that the presence of a central mental health authority increased the potential to ensure effective coordination. For example, Latin America and Caribbean countries have developed mental health units within the health ministry capable of overseeing coordination (PAHO, 2008). Coordination across countries in the initial phases of reform played a crucial role, by sharing technical support and experiences of implementation (PAHO, 2008). Authors highlighted the relevance of developing intersectoral coordination, which may act as a safety net for persons with serious mental health illness reducing acute episodes (PAHO, 2008).

Studies mentioned how increases in psychiatric population and fiscal strain on state mental hospitals drove governments to develop an alternative mental health strategy (McCubbin, 1994, Mechanic & Rochefort, 1990). The pressure on fiscal resources -partly linked to economic crisis-made the costs of mental health hospitals and their inefficiency more apparent (PAHO, 2008). Also, the direct transference of funds -from reduced hospital expenditure-to community-based services was mentioned as a factor that fostered the transference of patients from state hospitals to alternative placements in the community (Mechanic & Rochefort, 1990). Finally, the growth of disability insurance was understood as a facilitator of the process of discharging service users from psychiatric hospitals by contributing to their support in the community (Mechanic & Rochefort, 1990).

#### 2. Knowledge/Science

Interdisciplinary research focusing on the legal and economic factors which influence PDI processes and practices was valued (Mechanic & Rochefort, 1990; PAHO, 2008). The elucidation of adverse effects of institutions on individual patients (Anderson et al., 1998; Bennett & Morris, 1983; Kleiner & Drews, 1992; Mechanic & Rochefort, 1990) together with the documentation of human rights violations in mental health hospitals helped in catalysing the reform process (Bennett & Morris, 1983; PAHO, 2008). More generally, some authors stressed that conceptual clarity regarding the application of a biopsychosocial model to the mental health field (McCubbin, 1994) and the interpersonal aspect of mental health (Bennett & Morris, 1983; Kleiner & Drews, 1992) helped in the rolling up of the Deinstitutionalisation processes.

In the early stages of PDI in the USA, the allocation of research grants to state mental health hospitals developing pilot testing of outpatient treatment and rehabilitation helped in the shift of funds from mental hospitals into general hospitals (Weiss, 1990). The dissemination of early experiences of innovative policy implementation in mental health facilitated the adoption of Deinstitutionalisation practices in other regions (Shen & Snowden, 2014). Finally, the development of psychotropic medication and the reduction of psychiatric symptomatology helped to build trust in the implementation of less coercive management plans that were feasible to apply at the community level (Anderson et al., 1998; Bennett & Morris, 1983; Bredenberg, 1983; Freedman & Moran, 1984; Kleiner & Drews, 1992; Mechanic & Rochefort, 1990).

#### 3. Power, interests, and influences

This category points to the role of social movements and organizations in influencing the development of Deinstitutionalisation processes. This includes advocacy actions and legal transformations.

Mental health professional groups and civil society organizations were seen as key agents contributing to overcome stigma and change the delivery of mental health services (Weiss, 1990). Some authors emphasized the importance of promoting the active involvement of civil society groups (Oshima & Kuno, 2016). Finally, authors highlight how the internationalization of mental health reforms puts increasing pressure on other countries to jump on the “bandwagon” to avoid appearing antiquated (Shen & Snowden, 2014).

Recognition of the rights of people with disabilities and their defence by civil rights movements fostered the development of new mental health laws promoting less restrictive therapeutic alternatives and broader transformations on mental health systems (Anderson et al., 1998; Freedman & Moran, 1984; Mechanic & Rochefort, 1990; PAHO, 2008; Shen & Snowden, 2014). These changes involved expanding the supply of options in the community (Anderson et al., 1998; Freedman & Moran, 1984; Mechanic & Rochefort, 1990; PAHO, 2008) and relocating investment from institutions to community services (Swidler & Tauriello, 1995). In some countries, an extensive and strong network of community-based organizations provided opportunities for community participation, facilitating the effective integration of patients into the community (PAHO, 2008). This was accompanied by the divulgation of reports showing mistreatment of patients in hospitals, pushing public sensitivity against asylums (Anderson et al., 1998).

#### 4. Services and supports in the community

This category describes how the characteristics and distribution of community-based services and support for persons with SMI acted as facilitators in PDI processes.

Authors noted how policies around prevention in mental health, the integration of mental health services in primary health care centres (Kraudy et al., 1983; PAHO, 2008) and the accessibility of services (Mayston et al., 2016), together with social support such as supplementary income, can sustain community inclusion (Lamb & Goertzel, 1977), giving sustainability to Deinstitutionalisation. Adequate coordination across community-based services allowed the adequate externalization of users with complex needs (Cohen, 1983; Conway et al., 1994; Evans et al., 2012). Scaled up outpatient facilities including local acute hospitals and intermediate facilities (Abas et al., 2003; Bennett & Morris, 1983) were key in allowing mental health systems to reduce their reliance on inpatient care and limiting beds in psychiatric settings (PAHO, 2008). Plans to end seclusion and to support mental health professionals towards a transformation in their clinical practice were identified as a facilitator to the transition (Ash et al., 2015).

Other facilitators included the continuity of care after discharge (Sytema et al., 1996) and specific actions such as: developing mobile teams and home interventions as they facilitate access to service for users who can’t physically access needed services (John et al., 2010), mitigating self-stigma dynamics by allowing an active participation of users in their treatment through shared decision-making with professional staff (Chan & Mak, 2014; Matsea et al., 2019; Mayston et al., 2016) and supporting mechanisms for primary care workers such as a 24 hr hotline for assistance when it is required (Huang et al., 2017).

In terms of training, it is argued that a reform such as PDI requires the development of an educational infrastructure including local health training networks for continuing education and training needs, and targeting providers, service-users, volunteers, family members and others (Wasylenki & Goering, 1995). The incorporation of non-specialized, community-based workers trained on mental health prevention and promotion is also highlighted (Mayston et al., 2016).

Expanding user’s freedom to choose among service options was a central facilitator. This includes models of self-directed care, where users are given a budget to choose between service options (Kalisova et al., 2018). Experiences from the US, Germany and England show that patients used their budget to pay for care from their relatives, avoiding the use of institutionalized settings and preventive care options, thus shifting from crisis intervention to early interventions (Alakeson, 2010). Self-directed care improved user’s autonomy and has proved to be an effective preventive intervention (Alakeson, 2010).

#### 5. Workforce

Facilitators related to community mental health services workforce were organised under this category. Strategies around training and skills include enhancing psychiatric aspects in health curriculum and provision of grants to complete training and research projects. This attracted students from other professions to the community mental health field (Weiss, 1990). Having previous experience in general medicine before training into psychiatry appeared to support a culture of community-based work and a strong collaboration with primary care teams (PAHO, 2008).

#### 6. Exogenous factors

Factors indirectly affecting the feasibility of implementing Deinstitutionalisation policies were gathered under this category. This includes the role of exogenous shocks (e.g. conflict and humanitarian disasters) (Shen & Snowden, 2014) in bringing wider public attention to patients’ living conditions. A study also mentioned how the end of dictatorial regimes brought attention to human rights issues in psychiatric care, facilitating the process of Deinstitutionalisation in countries such as Argentina, Brazil and Chile (PAHO, 2008).

## Discussion

A marked decline in interest on psychiatric institutions across the global mental health literature has been noted by Cohen and Minas (2017) being absent from important prioritization exercises like the Grand Challenges in Global Mental Health (Collins et al. 2011). The authors argue that although establishing high-quality community mental health services is crucial for improving the lives of people with severe mental disorders, an exclusive focus scalability overlooks ongoing deficiencies in treatment quality and human rights protections in psychiatric institutions. Given their role in human rights abuses experienced by people with mental disorders, PDI efforts should receive more attention.

In response to this call, this article organised the available evidence around PDI, to assist in planning and conducting contextually relevant studies about and for the process. Drawing on the review, the following section introduces a set of proposals while reflecting on the limitations and problems with the available literature.

### Moving Psychiatric Deinstitutionalisation Forward

The transition from a system centred on long-term psychiatric hospital care to one centred on community-based services is complex, usually prolonged and requires adequate planning, sustained support and careful intersectoral coordination. The literature documenting and discussing PDI processes is vast, running across different time periods, regions, socio-political circumstances, and disciplines, and involving diverse models of institutional and community-based care. Based on this scoping review, we propose five key considerations for researchers and policymakers involved in PDI efforts:

1. *Needs assessment, design and scaling up*: An adequate assessment of the institutionalized population is required, to shape existing and new community-based services around their needs and preferences. An adequate analysis of the correlation of forces required to unlock institutional inertia is crucial.
2. *Financing the transition*: A comprehensive and sustainable investment is necessary, and the different aspects of the transition should be adequately costed, including new facilities, support of independent living, training, new professional roles, and the reinforcement of primary health care.
3. *Workforce development*: The workforce should be aligned with the transition from the outset. Elements such as training, incentives and guarantees of job stability are required. Curricular changes in psychiatric training, including more emphasis on community-based care and recovery-oriented practices, are necessary.
4. *PDI implementation*: The implementation process requires political resolve, careful monitoring, and an ability to respond to unexpected challenges. PDI represents a crucial learning opportunity for further scaling up.
5. *Monitoring and quality assurance:* Results of the process need to be carefully assessed against clear operational goals. The perspectives of users, caregivers, and the workforce should be incorporated into the assessments. The development of an assessment strategy detailing clear outcomes that incorporate financial and organisational dimensions is advised. Thorough documentation of PDI process, including achievements and setbacks should be done to build a reliable and diverse evidence-base for action.

A multifaceted strategy, clear and strong leadership, participation from diverse stakeholders and long-term political and financial commitment are basic elements in the planning of PDI processes. Nonetheless, implementation dynamically responds to local conditions, widely differing across countries and regions. What appears as a barrier or a facilitator can vary according to a specific context.

These findings are consistent with other reviews. In their review article on deinstitutionalization and the “home turn” from the 1990s, Hall et al., (2021) pose questions about the outcomes of the transition of care and the extent of social inclusion achieved. They argue that advancing the process of psychiatric deinstitutionalization and providing services and support to patients in their homes and demands significant financial investments and human resources. One crucial aspect is the capacity of front-line workers to promote the social inclusion of patients in the community. Advancing PDI processes necessitates sustained development, time, and support from all stakeholders.

### Limitations in the literature: Time, space, process, and voice

The literature on PDI is diverse, which makes synthesis endeavours difficult. Although promoted as a global standard in psychiatric and social care, the multiplicity of contexts in which the policy has been implemented limits the possibility of finding common ground. In their systematic review of the current evidence on mental health and psychosocial outcomes for individuals residing in mental health supported accommodation services, McPherson and colleagues (2018) noted how the variation in service models, the lack of definitional consistency, and poor reporting practices in the literature stymie the development of adequate synthesis.

Similarly, in a recent systematic review of psychiatric hospital reform in LMICs, Raja et al. (Raja et al., 2021 pp. 1355) expressed regret over the “dearth of research on mental hospital reform processes,” indicating how poor methodological quality and the existence of variation in approach and measured outcomes challenged the extrapolation of findings on the process or outcomes of reform. Of the 12 studies they selected, 9 of them were rated as weak according to their quality assessment.

Beyond the challenges posed to synthesis efforts and through conducting this review, we identified four wider problems affecting the literature documenting PDI planning and implementation. They are related to *time, location, focus*, and *voice*.

In terms of *time*, most of the work addressing PDI was developed at the end of the 1970s through the 1980s and early 1990s. After this, there are barriers and facilitators documented which indirectly relate to the development of community-based services and their evaluation, with PDI as the “background” but not as the main object of attention. Also, the date of the search -May 2020-could potentially exclude studies that worked with data from the pre-COVID period.

When it comes to *location*, while there is a wealth of literature on the topic, it is important to note that much of it is based on the experiences of the USA and Western Europe. The documentation of PDI in regions outside of the ‘global north’ is typically limited to personal testimonies from process leaders, which may lack systematicity and are usually published in languages other than English. This can restrict their accessibility and dissemination.

In terms of *focus*, most studies have a clinical orientation, evaluating various outcomes that are directly or indirectly related to PDI. However, the process itself, has received little attention. An exclusive emphasis on outcomes can obscure the administrative, legal, and political complexities of carryng out a psychiatric reform, this, hinder the dissemination of important lessons.

Finally, it’s worth noting that important *voices* are often missing from available studies and reports on PDI processes. While some studies do consider the experiences and engagement of caregivers, healthcare workers, and patients, they are still in the minority. This can create a skewed understanding of the impact of PDI, as these individuals play crucial roles in shaping the process and its outcomes. The same goes for the different communities where patients have developed their lives after PDI.

These limitations have significant consequences. It’s unclear whether the evidence extracted from experiences in high-income countries in North America and Europe can directly inform processes in other regions, including low- and middle-income countries (LMICs). While it’s possible to identify common pitfalls, barriers, and needs, this identification must be accompanied by up-to-date local research to ensure that the evidence is relevant and applicable to specific contexts.

The involvement of patients and communities affected by institutionalization in the design and implementation of research and policy should be central in a renewed PDI agenda. The recently launched Guidelines on deinstitutionalization, including in emergencies, by the United Nations Committee on the Rights of Persons with Disabilities represent a pioneering effort in this direction (OHCHR, 2022).

At the same time, qualitative and ethnographically oriented case studies are required to closely examine PDI efforts while remaining attentive to diversity and local creativity beyond global normative parameters of success and failure. Furthermore, reflexive, and flexible approaches to research synthesis are necessary to capture and assess the wealth of lessons learned from diverse engagements with deinstitutionalization across the globe.

This article offers a preliminary and general classification of barriers and facilitators that can inform the development of relevant research through various methodologies and other literature. The categories can be modified and customized based on the evidence from various settings. As far as we know, this classification is not yet present in the existing literatura

## Conclusion

Institutional models of care continue to dominate mental health service provision and financing in many countries, leading to a continued denial of the right to freedom and a life in the community for individuals with mental health conditions and associated disabilities. The successful implementation of PDI requires detailed planning, sustained support and coordinated action across different sectors.

This review identifies the factors impacting PDI processes, according to the available literature. Barriers and facilitators are organised in fifteen thematic groups. The results reveal that PDI processes are complex and multifaceted, requiring detailed planning and commensurate financial and political support. We have offered five considerations for policymakers and researchers interested and/or involved in PDI efforts.

There are many lessons to be learned from the processes described in the literature, and many areas where research has been insufficient. Barriers and facilitators will differ in response to the legal, institutional, and political characteristics of each region and country. This categorization can be adapted to national realities and different levels of policy progress in PDI, to guide research and policy efforts. We call for methodological innovation and the involvement of affected communities as key elements of a renewed research agenda around this neglected aspect of mental health reform worldwide.

## Supporting information

(1) Supplementary Material A Protocol

(2) Supplementary material B PRISMA-ScR-Fillable-Checklist

(3) Supplementary Material C Tailored search strategy to different databases

(4) Table 1 Inclusion and Exclusion criteria

(5) Table 2 Data extraction form

(6) Supplementary Material D Data coding process example

(9) Table 3 Summary characteristics of included studies

(10) Table 4 Studies characteristics

(11) Table 5 Barriers

(12) Table 6 Facilitators

## Data Availability

All data produced in the present study are available upon reasonable request to the authors.

## Required Manuscript Parts

### Author Contribution statement

M.I and M.C conceived the idea for the project. J.U and C.M developed the framework to conduct the systematic search, which J.G performed. J.U and J.G, established the eligibility of articles under the supervision and with the contribution of C.M. J.G and J.U extracted the data of the selected articles. J.G, J.U and C.M coded the article contents and created the categories iteratively through rounds of revision and adjustment. J.U and C.M produced an early draft of the manuscript.

F.T reviewed several versions of the manuscript. The final manuscript was discussed and improved by all the authors. C.M and J.U coordinated the development of the manuscript.

### Conflict of Interest statement

Conflicts of Interest: None

### Data Availability Statement

The authors confirm that the data supporting the findings of this study are available within the article [and/or its supplementary materials.]

## Notes

### Competing Interest Statement

The authors have declared no competing interest.

### Funding Statement

The work reported in the article has been supported by the Chilean National Agency for Research and Development (ANID), Initiation FONDECYT N. 11191019, given to Cristian Montenegro.

