## Supplementary material for "Moving psychiatric deinstitutionalisation forward: A scoping review of barriers and facilitators": (1) Supplementary Material A Protocol

**Scoping Review Protocol**

Prepared for Registration to Open Science Framework

Submitted on 2^nd^ of July of 2020

**Title:** Barriers and Facilitators involved in the Process of Psychiatric Deinstitutionalization: A Scoping Review

**Author Names:** Jorge Urrutia Ortiz^a^, Cristian Montenegro Cortés^b,d^, Matías Irarrazaval Dominguez^b^, Josefa González Moller^c^

**Affiliation and contact details for the corresponding author:**

^a^ Psychology Department, University of Chile, Santiago, Chile.

^b^ Department of Mental Health, Division of Prevention and Control of Diseases, Chilean Ministry of Health, Santiago, Chile.

^c^ Child and Adolescent Mental Health Services, Centro de Referencia de Salud El Pino, Santiago, Chile.

^d^ School of Nursing, Pontificia Universidad Católica de Chile, Santiago, Chile.

**Scoping Review Objective:** This study will examine the extent and range of available research regarding barriers and facilitators involved in the process of deinstitutionalization of adults with mental health difficulties in order to improve understanding of this process and propose policy actions to advance this process.

**Scoping Review Question:**

What is known about the barriers to, and facilitators of, deinstitutionalization processes for people with mental illness?

**Background:**

Psychiatric deinstitutionalization (DI) is a complex process that started in Europe and North America during the 1940´and 1950´. More than simply involving the closure of long-stay facilities, it constitutes an active socio-political movement with two broad components: (i) the reduction of traditional institutional care of people with mental health difficulties and (ii) the development of comprehensive community-based mental health services aiming to promote social inclusion and full citizenship (Bachrach, 1978; Rotelli et al., 1987; Saraceno, 2003).

Several historical and social forces were involved on the development of DI including a growing awareness of the poor conditions of inmates, and dehumanizing effects of prolonged institutionalization, the high cost of maintaining long-stay mental health institutions, as well as the introduction of psychotropic medication (Taylor Salisbury et al., 2016; Turner, 2004; Yohanna, 2013). Advocacy and human rights movements also encouraged this transformation, looking to destigmatize mental health difficulties and improve the quality of mental health care (Hillman, 2005; Mezzina et al., 2019).

After World War II many Western countries started to explore alternatives to psychiatric hospitals and a significant proportion of institutionalized people were discharged from inpatient units to find an heterogeneous degree of support in community settings (Grob, 2014; Turner, 2004; Yohanna, 2013). It is important to acknowledge that this process is not free of risks, as the Esidimeni tragedy in South Africa recently showed (Moseneke, 2018). The implementation of ID cannot result in the abandonment of vulnerable populations and for this it is necessary to promote a geographical decentralisation of resources, developing community mental health services that support primary care workers and inviting community members who have mental disorders and their family members to participate in advocacy and service delivery (Capri & Swartz, 2018; Benedetto Saraceno et al., 2007).

Currently, deinstitutionalization is a significant component of national mental health policies and it is encouraged by international agencies (Shen & Snowden, 2014). The World Health Organization (WHO) (WHO, 2013) and the Movement for Global Mental Health (Campbell & Burgess, 2012; Thornicroft et al., 2016) have both stated that mental health care should be shifted from hospital to community-based treatment facilities. Notably, the WHO Initiative for Mental Health proposes to scale up interventions and services across community-based settings as one of the main strategies to achieve universal health coverage by 2023 (WHO, 2019).

Despite this general consensus and encouragement, DI has shown to be a highly complex whose progress and scope greatly varies across countries (Goldman et al., 1982; Hudson, 2019). As stated in a milestone report by the WHO and the Gulbenkian Global Mental Health Platform: “*despite decades of promoting deinstitutionalization and community-based care, mental hospital-based care still dominates service delivery in most countries. (…) [M]ost countries continue to spend the vast majority of their scarce resources on the inefficient and frequently inhumane approach of managing few people with mental disorders exclusively in long-stay institutions¨* (2014, p. 12)

For processes of deinstitutionalization to start or to move forward at a larger scale, it is necessary to understand the main factors both delaying and promoting DI. Despite the extensive body of literature on DI, no previous attempts to apply a systematic methodology to explore its barriers and facilitators where found. This study will examine the extent and range of available research regarding barriers and facilitators involved in the process of deinstitutionalization of adults with mental health difficulties in an attempt to improve understanding of this process and propose policy actions to advance this process. Given that the body of literature on this topic has not been comprehensively reviewed, the Scoping Review (ScR) (Arksey & O’Malley, 2005) was considered the appropriate methodology. The goal of ScR is “*to map rapidly the key concepts underpinning a research area and the main sources and types of evidence available* (…) *especially where an area is complex or has not been reviewed comprehensively before”*(Mays et al., 2001)*.*

For this review, a barrier was defined as any factor that limits or restricts the progress of deinstitutionalization at different stages of the process and across different stakeholders’ levels. This may include, but is not limited to, issues related to public-health priority agenda (Shen & Snowden, 2014); challenges in the implementation of mental health care in community settings (Kormann, R. J., & Petronko, 2004; Benedetto Saraceno et al., 2007) and communities’ acceptability and the presence of stigma (Fisher et al., 2005; O’Doherty et al., 2016).

Correspondingly, a facilitator was defined as any factor that fosters or enhance this process. Positive factors or facilitators that promote mental health reforms towards deinstitutionalization can be such as the presence of well-organized social activism supporting the rights persons with mental health problems (Anderson et al., 1998), the acceptance of mental illness as a human condition (Gostin, 2008), social paradigms that enhance social inclusion and citizenship (Fakhoury & Priebe, 2002; B. Saraceno, 2003) and political willingness (Benedetto Saraceno et al., 2007). Because we focused on barriers and facilitators that could be modified by an intervention, we did not consider age, sex or ethnic background as barriers or facilitators.

The synthesis of this information will allow us to establish a list of suggestions on ways to move forward, differentiated between country profiles according to their degree of progress in ID processes.

**Inclusion Criteria:**

**Types of participants:** Studies focused on adult users of long-term (more than 60 days) mental health services. Studies meeting the above criteria but where participants had a background of a long-term stay in Children Services facilities (children ward, orphans’ asylum, group home or residency) will be excluded in the light of the potential differences that may affect the process of deinstitutionalization of Mental Health organizations from Social Services.

**Concept:** Studies focused on providers, caregivers (family/friends) and users’ account on barriers and facilitators of the psychiatric deinstitutionalization process.

**Context:** Studies conducted in a community mental health setting (including Day Service Units).

**Types of sources:** papers (primary studies, textual papers, technical and governmental reports, calls to action, theoretical and political discussions, historical studies, book chapters and reviews) both published and unpublished (grey literature) wrote in English or Spanish.

**Search Strategy:** Studies will be identified by searching three electronic databases, Medline, CINAHL and Sociological Abstracts. Further information will be obtained through screening reference lists, contacting experts in the field and searching grey literature repository and library catalogues: PsycExtra, OpenGrey and New York Medical Academy.

The search strategy (Table 1) is informed by previously published systematic reviews on adults with severe mental health impairment (Lean et al., 2019; Richardson et al., 2019), barriers and facilitators to healthcare access (Adauy et al., 2013) and deinstitutionalization process (May et al., 2019).

| Category | Key Terms |
| --- | --- |
| Population | ((disorder OR disease OR Illness OR problem OR disability OR condition) NEAR/2 (Severe OR Serious OR chronic OR enduring) NEAR/2 (Mental OR Psychopatolog* OR psychiatric OR schizo* OR psycho?i* OR hebephreni* OR oligophreni*)) |
| Concept | (((("health care"[All Fields] OR "healthcare"[All Fields]) AND ("research"[All Fields] OR "evaluation"[All Fields] OR "utilization"[All Fields] OR "availability"[All Fields] OR "access"[All Fields] OR "accessibility"[All Fields] OR "coverage"[All Fields] OR "barrier"[All Fields] OR "barriers"[All Fields] OR “difficult*” OR "facilitator"[All Fields] OR "facilitators"[All Fields] OR "usage"[All Fields] OR "intake"[All Fields])) OR "health services accessibility"[MeSH Terms]) OR "delivery of health care"[MeSH Terms]) OR "health services needs and demand"[MeSH Terms] |
| Context | (Deinstitutionali?ation or Community or "independent living" or noninstitution* or "step?down facility" or "supported living" or “transition” or “transfer” or “ hand?off” or “hand?over” or “discharge”) |

All references obtained through the electronic database search and hand searching will be pooled in EndNote reference manager software. Pilot-testing (with three reviewers) will be applied to 50 citations in batches until obtaining a minimum of 80% of agreement across reviewers. Initial eligibility will be assessed by JU and JG based on title and abstract and full texts of the selected articles screened. Differences in opinions will be discussed and a final decision on their eligibility will be arrived at after discussion with CM.

**Extraction of the results or “charting the results”:** A specific data extraction form will be created to record full study details and guide decisions about the relevance of individual studies to the review questions. After piloting and revising the form, a Microsoft Excel spreadsheet will be populated with the following information: authors, year of publication; location of study; aims; study design; study population and sample size; setting; provider type; barriers and facilitators related to the process of deinstitutionalization. Data will be independently extracted by two reviewers.

**Presentation of the results:** Given that ScR methodology is presented as a first attempt to map the state of research on the topic, (Arksey & O’Malley, 2005; Peters et al., 2015), no attempts will be made to assess the methodological quality of the included studies.

Data will be analysed using QSR’s NVivo 11. Thematic Synthesis (Harden, 2010; Lucas et al., 2007; Thomas & Harden, 2008) following a three-stage process will be applied. Firstly, encompasses free coding the themes described in the extracted data. Secondly, organizing the codes into related areas of “descriptive themes” by looking for similarities and differences between and within them. Finally, three reviewers will independently examine the descriptive themes and their associated data in the light of the review question to infer barriers, facilitators, and implied recommendations for future research on the process of deinstitutionalization.
