## Supplementary material for "Moving psychiatric deinstitutionalisation forward: A scoping review of barriers and facilitators": (3) Supplementary Material C Tailored search strategy to different databases

**Supplementary Material C:** Tailored MESH terms to each database

|  | MESH terms/controlled vocabulary) | | |
| --- | --- | --- | --- |
| Dimension | **Medline (OVID)** | **CINAHL (EBSCO)** | **Sociological Abstracts (PROQUEST)** |
| Population | Schizophrenia/ or affective disorders/ or paranoid disorders/ or psychotic disorders/  Mental Disorders/ | (MH "Mental Disorders") OR  Schizophrenia | "psychosis/psychoses/ psychotic/ psychotics" OR "schizophrenia" OR "mental disorders" |
| Concept | Health Services Accessibility/  "Delivery of Health Care"/ | MH "Health Services Accessibility") OR (MH "Health Care Delivery") OR (MH "Health Services Needs and Demand") | Health Services Accessibility/  "Delivery of Health Care"/  health care utilization/ or health care delivery/ |
| Context | Deinstitutionalization/  Community Mental Health Services/  Hospitals, Psychiatric/  NOT (Child welfare/ or Orphanages/) | "Hospitals, Community") OR (MH "Psychiatric Home Care") OR (MH "Community Mental Health Services") OR (MH "Community Health Centers") OR (MH "Hospitals, Psychiatric") NOT Child welfare | "hospitals, psychiatric" OR "deinstitutionalization" OR "community mental health" |
