## Supplementary material for "Moving psychiatric deinstitutionalisation forward: A scoping review of barriers and facilitators": (4) Table 1 Inclusion and Exclusion criteria

| Table 1: Inclusion and Exclusion criteria | | |
| --- | --- | --- |
|  | **Included** | **Excluded** |
| Population | - Studies focused on adult users of long-term mental health services (stays longer than 60 days). | - Studies meeting the above criteria but where participants had a background of a long-term stay in Children Services facilities (children ward, orphans’ asylum, group home or residency) or specialized substance abuse services. |
| Concept | Studies focused on providers, caregivers (family/friends) and users’ account on barriers and facilitators of the psychiatric deinstitutionalization process. Studies focused on PDI processes were included regardless of the study aims. Studies focused on reporting outcome measures related with the community mental health system where only included if they involved a reform process in the context of PDI. | - No mention of any facilitator or barrier related to the process of PDI.  - Studies where the researchers could infer the presence of a barrier o facilitator of PDI but no direct link with PDI processes were clearly set out by the authors were excluded. |
| Context | - Studies conducted in mental health setting.  - No restrictions were placed on the location of intervention delivery (i.e. hospital, day services, community health centre, homes). | - No description of the mental health services provided |
| Type of Source | Published and unpublished (grey literature) sources including primary studies, textual papers, technical and governmental reports, calls to action, theoretical and political discussions, historical studies, book chapters and reviews. |  |
| Language | - Studies wrote in English or Spanish | - All other languages. |

* In the light of the potential differences that may affect the process of deinstitutionalization of Mental Health organizations from Social Services and Specialized Substance Abuse Services (like penal law involvement), this kind of interventions will be excluded.
