## Supplementary material for "Moving psychiatric deinstitutionalisation forward: A scoping review of barriers and facilitators": (5) Table 2 Data extraction form

| Table 2: Data Extraction Form | |
| --- | --- |
| Study Information | Correspondence Author |
|  | Title |
|  | Year of Publication |
|  | Country in which the study was conducted |
|  | Aim of study |
|  | Study Design |
|  | Population description |
|  | Nº of participants |
|  | Setting |
|  | Provider type |
| Outcomes | Barriers to Psychiatric Deinstitutionalization |
|  | Facilitators to Psychiatric Deinstitutionalization |
|  | Policy Advice |
