## Supplementary material for "Moving psychiatric deinstitutionalisation forward: A scoping review of barriers and facilitators": (6) Supplementary Material D Data coding process example

**Table: Data coding process example**

|  | **Inductively created codes** | | **Data extracted from primary studies** |
| --- | --- | --- | --- |
|  | **Terciary construct** | **Secondary construct** | **Primary construct** |
| Facilitator | Power, interests and influences | Advocacy from professional organizations/groups | *“I have already mentioned the associations of mental health professionals, some state mental health officials, and the organizations who spoke for mental patients and their families. The professional groups contributed as lobbyists, as educators and as bridges to the public who needed to overcome the stigma attached to mental illness. The Veterans Administration also contributed to change in the delivery of mental health services”* (Weiss, 1990) |
|  |  |  | "*Political skill was considered important because of the rigid attitudes and resistance to change among several groups, including health workers.(especially those working in the institutions subject to down-sizing), government officials, community members, and families and patients themselves. Writing about challenges with family members, Gurudatt Kundapurkar of India stated that, ‘[In rural areas], if the affected person is not able to contribute to family income/household chores they may admit her to the government hospital, invariably located at far off district headquarters, even with fictitious residential address so that the hospital will not be able to send her back home later. Fear of stigma is also one other reason for not taking recovered persons back home. Hundreds of such stable persons with mental illness are stuck in these institutions for years’”* (WHO and the Gulbenkian GMHP, 2014) |
|  |  | Involvement of families | *"The community mental health system would function better if it reduced family burden and involved families in long-term care planning for their relatives with SMI*"(Oshima & Kuno, 2006) |
|  |  |  | "*the 2005–2010 Mental Health Action Plan (MHAP), which promotes public access to care, continuity of services, quality of life, effectiveness and efficiency in the health care system, and the hierarchization of care. Its goal is the development of community services that are readily accessible to the entire population, supported by the judicious use of second-line (specialized) and third-line (ultraspecialized) services."* (...) *MHAP encourages mental health professionals to be more open toward families. They also pointed to an evolution in mental health practice reflected in a more comprehensive view of patients that extends to their environment and, thus, their families."* (Lavoie-Tremblay et al., 2012) |
|  |  | Service users movements and demands | "*Consumer movements (...) became important advocates for expanded and improved services and spurred development of empowering self-help programs and consumer operated services."*(Rosenheck, 2000) |
|  |  |  | *"Recent decades have seen a dramatic reduction in the mental hospital population of the United States. The resident population in State and County mental hospitals declined from 559,000 in 1955 to 191,000 in 1975 to 120,000 in 1985. This phenomenon has been variously attributed to the development of effective psychotropic medications, to the social psychiatry movement’s stress on interpersonal etiologies and treatments, to the demonstrated anti-therapeutic effect of large mental hospitals, and to the desire of the Federal and State governments to control soaring costs of psychiatric hospitalization."*(Kleiner & Drews, 1992) |
