## Supplementary material for "Moving psychiatric deinstitutionalisation forward: A scoping review of barriers and facilitators": (9) Table 3 Summary characteristics of included studies

| Table 3: Summary characteristics of included studies | | Nº of studies |
| --- | --- | --- |
| Setting | Community mental health | 19 |
|  | Mixed | 15 |
|  | Inpatient | 10 |
|  | Residency | 4 |
|  | Primary care centre | 2 |
|  | Day Service | 1 |
|  | Emergency Department | 1 |
| Provider | Public | 30 |
|  | Other | 16 |
|  | Private | 5 |
|  | NGO | 1 |
| Language | English | 52 |
|  | Spanish | 0 |
