## Supplementary material for "Moving psychiatric deinstitutionalisation forward: A scoping review of barriers and facilitators": (10) Table 4 Studies characteristics

| Table 4: Study Characteristics of Included Studies | | | | | | | |
| --- | --- | --- | --- | --- | --- | --- | --- |
| First author and year | Country | Aim of study | Setting | Study Design | Type of end user or participant | Number of participants | Provider Type |
| Abas 2003 | New Zealand | To describe reasons for admission and alternatives to admission in a government funded acute inpatient unit | Inpatient Unit | Mixed Methods | Adult patients admitted to a psychiatric hospitalization in the South Auckland Health Mental Health Services | 255 admissions to an acute psychiatric unit in Auckland | Public |
| Aggett 2005 | UK | To describe the work of a busy Community Mental Health Team with outreach clients. Barriers to collaborative work and some of the team’s strategies to overcome them are delineated. | CMHC | Case series | Difficult to engage adult clients between 35 and 52 years old of a outreach community mental health team in an East London borough. | 4 service users | Public |
| Alakeson 2010 | USA | To examine a range of innovative self-directed care programs in England, Germany, the Netherlands, and the United States. | CMHC | Narrative style | Home and community-based long-term care service users with physical and cognitive disabilities | Inapplicable | Public |
| Allen 2007 | UK | To investigate predictors for out of area placements for people with challenging behaviors and also reports on their costs and basic characteristics. | Mixed | Descriptive Transversal | All people attending to services supporting children and adults with intellectual disability in a large area of South Wales in conjunction with health, education, unitary authority, voluntary and private sector commissioners and providers. | 1458 people | Public |
| Anderson 1998 | USA | To show the changes over 30 years in state institutional populations, interstate variability, movement of individuals into and out of state institutions, costs of state institutional care, and state institution closure as a result of social policy. | Inpatient Unit | Descriptive Longitudinal | Patients in institutions for mental disabilities and epileptics between 1950 and 1968 | Inapplicable | Public |
| Ash 2015 | Australia | To describe the implementation of recovery-based practice into a psychiatric intensive care unit, and report change in seclusion rates over the period when these changes were introduced (2011–2013). | Inpatient Unit | Mixed Methods | Consumers (average age 38 years) detained under the SA Mental Health Act. Eleven percent had been charged with or convicted of an offence with a custodial sentence. Common diagnoses were schizophrenia (32%), drug-induced psychosis (18%) and bipolar disorder (manic) (18%). The average length of stay was 11.5 days. | 63 people | Public |
| Barry 2002 | USA | To examine the relationship between age, the use of health services and level of functioning in patients with schizophrenia across the adult lifespan | Mixed | Descriptive Transversal | Veterans with schizophrenia drawn from the VA National Psychosis Registry who received a diagnosis of schizophrenia during a VA clinical encounter between 1999 and 2000. | 102.256 | Other: not described |
| Barton 1983 | USA | To discuss the role of mental hospital in the health care system for the elderly | Inpatient Unit | Narrative style | Inapplicable | Inapplicable | Mixed |
| Bennett 1982 | UK | To describe factors that fostered the deinstitutionalization process in the UK and its consequences in psychiatric services. | Mixed | Narrative style | Inapplicable | Inapplicable | Public |
| Bredenberg 1983 | USA | To present available documentation regarding the implications of residential integration of geriatric ex-mental patients and the well elderly and make recommendations for future action. | Residency | Narrative style | elderly discharged mental health service users | Inapplicable | Other: not described |
| Bryant 2004 | UK | To identify how the experience of attending day services met the needs of people with enduring mental health problems | Day Unit | Thematic analysis | patient population. | 39 people | Public |
| Chakraborty 2011 | UK | To compare measures of perceived racism, medication adherence and hospital admission between African- Caribbean and white British patients with psychosis | Mixed | Cohort study | participants aged 18–65 years; with a self-assigned ethnicity of Caribbean origin with either parents or grandparents born in the Caribbean; having a Research Diagnostic Criteria-defined psychotic symptom and in receipt of psychiatric services in north London, UK | 110 people | Public |
| Chan 2014 | Hong Kong | To examine the mediating role of self-stigma and unmet needs in the relationship between psychiatric symptom severity and subjective quality of life. | CMHC | Case series | Adults with schizophrenia spectrum disorders attending community mental health services in Hong Kong | 400 | Nonprofit organization |
| Chopra 2011 | Australia | To assess the long-term outcomes for the original cohort of 18 residents of the Footbridge Community Care Unit (CCU), a residential psychiatric rehabilitation unit at St Vincent’s Mental Health Melbourne. | ED | Cohort study | 14 schizophrenic and 4 people with schizoaffective disorder | 18 | Public |
| Cohen 1983 | USA | To clarify conceptions about mental illness in later life and promote the development of mental health services for the elderly in the community. | Residency | Narrative style | senior people with mental health difficulties living in housing arrangements | Inapplicable | Other: not described |
| Conway 1994 | UK | To report outcomes of community mental health services for people with schizophrenia who had shown very low levels of supported housing and structured day activity. | CMHC | Cohort study | patients from West Lambeth, London originally aged 20-65 years who satisfied the research diagnostic criteria for schizophrenia. | 51 people | Other: not described |
| Dorwart 1991 | USA | To assess the effect of changes in ownership and types of inpatient settings on the structure of the mental health services system. | Inpatient Unit | Analytic transversal | All nonfederal psychiatric hospitals in the United States, including community mental health centers with inpatient units between October 1987 and May 1988 | 915 hospitals | Mixed |
| Evans 2012 | USA | To describe the conversion of partial hospitals into recovery-oriented programs as part of system transformation. | CMHC | Narrative style | Stakeholders involved in a transformation of mental health service in a hospital | Inapplicable | Other: not described |
| Fakhoury and Priebe 2002 | UK | To provide an international overview of deinstitutionalization and review related issues as discussed in the current literature. | Mixed | Narrative style | Inapplicable | Inapplicable | Mixed |
| Freedman 1984 | USA | To identify and discuss the major policy issues related to the care of the chronically mentally ill, specifically the effects and implications of deinstitutionalization for this particular population. | CMHC | Case report | a 32-year-old schizophrenic who has spent more than 10 years in mental health institutions | Inapplicable | Public |
| Goering 1984 | Canada | To describe the 6-month and 2-year postdischarge outcome in each of five aftercare components for 505 subjects in a traditional system of service delivery. | Inpatient Unit | Cohort study | Adult people discharged from inpatient units in Toronto | 505 participants | Public |
| Grabowski 2009 | USA | To estimate the cross-state variation in the proportion of nursing home admissions indicating a mental illness, and the proportion of persons with mental illness admitted to nursing homes. | Residency | Descriptive Transversal | Nursing home admissions in the USA during 2005 | 1.150.734 new admissions | Private |
| Huang 2017 | Singapore | To design a general practitioner– partnership programme in an institute in Singapore to facilitate the transition to community services and gauge the impact of the interventions chosen to improve uptake of referrals. | CMHC | Mixed Methods | Stable mental health service users referred to the GP from December 2014 to January 2016 partnership programme in a mental health institute in Singapore | 238 service users | Private |
| John 2010 | India | To describe the successful management of a person with schizophrenia in the community through a primary care team in liaison with psychiatrist services. | CMHC | Case report | adult with psychotic symptoms living in an urban area of India | 1 person | Public |
| Kaffman 1996 | Israel | To report on an alternative community care program that has been developed and implemented in the Kibbutz Clinic for the treatment and rehabilitation of the severely mentally ill. | CMHC | Mixed Methods | adult people with a severe mental illness with poor functioning who participated in the program conducted in Telem, Israel, for at least 18 months and followed up for a minimum of 4 years. | 124 patients | Private |
| Kalisova 2018 | Czech Republic | To assess the effect of the S.U.P.R. psychosocial rehabilitation programme on the quality of care at the longer-term inpatient psychiatric departments | Inpatient Unit | Experimental not randomized (‘before and after’ design) | All Czech psychiatric hospitals focused on longer term inpatients, mainly with a diagnosis of schizophrenia. | 14 units for 499 patients with severe mental illness with complex needs | Other: not described |
| Kam-shing 2006 | China | To review and evaluate the implementation of community mental health in the People’s Republic of China | CMHC | Narrative style | Inapplicable | Inapplicable | Public |
| Kleiner 1992 | USA and Norway | To describe the experiences in the creation of innovative service delivery system which integrates psychiatric services with lay community support systems and patient social networks. | CMHC | Narrative style | Psychotic patients who had more than two years of cumulative hospitalization, and who could not be placed with relatives. | Inapplicable | Other: not described |
| Kraudy 1987 | Nicaragua | To assess the extent to which the new proposed model had been translated into a different way of delivering psychiatric care in Nicaragua | Primary Care Centre | Descriptive Transversal | children and adult patients attending one of the surveyed services for the first time irrespective of whether or not they had a psychiatric history. | 342 patients | Public |
| Lamb 1977 | USA | To assess the career of psychiatrically disabled people in the community | CMHC | Descriptive Transversal | Long-term psychiatrically disable people between 18 and 64 years old who live in the community in California with a psychotic diagnoses | 99 people | Private |
| Lavoie-Tremblay 2012 | Canada | To describe how families and decision makers perceive collaboration in the context of a major transformation of mental health services and to identify the factors that facilitate and hinder family collaboration. | CMHC | Thamatic Analysis | family members of users of mental health services and key decision makers on the mental health service | 54 family members and 22 decision makers | Public |
| Mallik 1998 | USA | To identify perceived barriers to community integration in people with psychiatric disabilities, in the areas of skills, environmental support, and community resources. | Inpatient Unit | Case series | People with psychiatric disabilities in the Alliance of Psychiatric Rehabilitation Program in Baltimore County, Maryland | 42 people | Public |
| Manuel 2012 | USA | To explore the experience of women with severe mental illness in transition from psychiatric hospital care to the community. | Residency | Thematic analysis | women living in transitional residences on the grounds of two state-operated psychiatric hospitals in the New York City metropolitan area, awaiting discharge to both supervised and independent housing in New York City. | 25 women | Public |
| Matsea 2019 | South Africa | To explore the views of different stakeholders about their roles as support systems for people with mental illness and their families in a rural setting. | CMHC | Content Analysis | Stakeholders comprising traditional health practitioners (faith and traditional healer), traditional leaders, church members, home-based care team and police officers from Mashashane, a rural setting in Limpopo Province, South Africa | 41 stakeholders | Public |
| Mayston 2016 | Ethiopia | To engage key stakeholders in participatory planning for a shift to mental health care integrated into primary care, and to explore their perspectives on acceptability and feasibility of the change. | CMHC | Framework analysis | key stakeholders (healthcare administrators and providers, caregivers, service-users and community leaders) living in Butajira town | 11 service users, 27 caregivers, 15 health extension worker and 10 health center workers | Public |
| McCubbin 1994 | Canada | To reevaluate the recent tendency to attribute economic causes to deinstitutionalization and its subsequent "treatment in the community" mental health systems | Mixed | Narrative style | Inapplicable | Inapplicable | Mixed |
| Mechanic 1990 | USA | To provide a comprehensive overview of the causes, nature, and consequences of the practice of deinstitutionalization in the Unites States | Mixed | Narrative style | Inapplicable | Inapplicable | Mixed |
| O'Doherty 2016 | Ireland | To document the views of family members of people with an intellectual disability regarding implementation of a personalized model of social support in Ireland. | CMHC | Grounded theory | parent, adult sibling or extended family member of a person receiving full-time residential supports from the agency | 40 family members | Public |
| Oshima 2006 | Japan | To explore how the introduction of community-based care has changed the role of psychiatric hospitals and families in caring for people with mental illness by examining the different types of living settings of clients treated for schizophrenia in Kawasaki as compared with a similar group of clients nationally | CMHC | Descriptive Transversal | adults with a diagnosis of schizophrenia living in the community and hospitalized in Kawasaki and the rest of Japan | 3.845 people living in Kawasaki and 448.000 living in Japan | Private |
| PAHO 2008 | Argentina, Brazil, Chile, Cuba, Jamaica and Mexico | To convey some of the more innovative experiences to reform mental health services implemented in Latin America and the Caribbean | Mixed | Narrative style | Inapplicable | Inapplicable | Public |
| Rizzardo 1986 | Italy | To analyse the impact of the reform on health care delivery by the general practitioner in an urban district in the Veneto region. | Primary  Care Centre | Descriptive Transversal | General practitioners working in a psychiatric service run by the University of Padua by 1983 | 24 general practitioners | Other: University facilities |
| Rose 1979 | USA | To analyse deinstitutionalization policy on the sector of community mental health care and review its accomplishments and difficulties. | Mixed | Narrative style | Inapplicable | Inapplicable | Mixed |
| Rosenheck 2000 | USA | To review the relationship between mental health service delivery and the community in which it is embedded. | CMHC | Narrative style | Inapplicable | Inapplicable | Mixed |
| Schmidt 2000 | Canada | To examine how psychiatric rehabilitation fits within a remote First Nations community. | CMHC | Thematic analysis | service providers, consumers and family members of aboriginal people with severe mental illness living in northern British Columbia. | 10 stakeholders | Public |
| Semke 1999 | USA | To explore system outcomes of interventions that were aimed at lowering high use of long stay state hospitals. | Mixed | Descriptive Transversal | adults living in the Washington state who experienced one psychiatric hospitalization of 30 days or more, or three or more psychiatric hospital admissions during a "pre-reform" period (1988) or after implementation of reform interventions (between 1991 and 1993) | 2.646.307 high utilizers of state hospitals | Public |
| Shen and Snowden 2014 | USA | To examine whether the institutionalization of deinstitutionalization policy changed the supply of psychiatric beds in 193 countries from 2001 to 2011. | Inpatient Units | Echological study | Mental health systems as units | 193 countries | Public |
| Stelovich 1979 | USA | To describe factors related to deinstitutionalization leading to transfer mental health service delivery from civil mental health hospitals to prison facilities. | Inpatient Unit | Narrative style | Psychiatric patients transferred to prison facilities in Massachusetts | Inapplicable | Public |
| Swidler 1995 | USA | To describe the political processes leading to the Community Mental Health Reinvestment Act passage, the obstacle overcome by legislative negotiators and implementation issues. | Mixed | Narrative style | Inapplicable | Inapplicable | Public |
| Sytema 1996 | Italy and Netherlands | To compare the treatment of severely mentally ill patients in a community mental health service without the back-up of a mental hospital with the treatment provided in an institution-based system in which mental hospital are still predominant. | Mixed | Cohort study | Patient with schizophrenia that contacted a service at least once in 1988 or in 1989 in Groningen (The Netherlands) or South-Verona (Italy) | 812 patients | Mixed |
| Wasylenki 1995 | Canada | To describe the authors' involvement in three service delivery projects in Ontario and discuss how, by assuming multiple roles, they were able to ensure that planning and policy development were informed by current knowledge. | Mixed | Narrative style | Inapplicable | Inapplicable | Public |
| Weiss 1990 | USA | To analyze deinstitutionalization policies implemented in 1946 and 1963 in USA | Mixed | Narrative style | Inapplicable | Inapplicable | Public |
| WHO 2014 | Malaysia, Japan, Ethiopia, Brazil, Nigeria, Uganda, UK, Iran, Italy, Portugal, Cambodia, Philippines, Spain, New Zealand, Usa, Sri Lanka, Chile, India, Republic of Korea, The Netherlandands, Zambia, Indonesia, Tanzania, Singapore, Lithuania, Australia, Georgia, Vietnam, South Africa, Ghana, Sweden | To capture lessons learnt from those who have been involved directly with deinstitutionalization and/or expanding community-based services and identify innovative strategies and methods associated with success of this process. | Mixed | Mixed Methods | mental health experts involved directly with deinstitutionalization and/or expanding community-based services | 78 people | Public |

*CMHC: Community Mental Health Centre; ED: Emergency Department
