## Supplementary material for "Moving psychiatric deinstitutionalisation forward: A scoping review of barriers and facilitators": (11) Table 5 Barriers

**Table 5: Barriers to the process of psychiatric deinstitutionalization**

| **Category** | **Descriptive themes** | **References** |
| --- | --- | --- |
| 1. Planning, leadership, and funding | Mental Health Policy: Responsibility/accountability | Rose, 1979 |
|  | Reform fragility: charismatic leadership | PAHO, 2008 |
|  | Reform fragility: Lack of synchronization between bed reduction and development of CBMHSs | Freedman & Moran, 1984; Shen & Snowden, 2014 |
|  | Reform fragility: Unaccountability of failure | Freedman & Moran, 1984; Rose, 1979; Rosenheck, 2000 |
|  | Funding: Continuity of community care | McCubbin, 1994; Mechanic & Rochefort, 1990; PAHO, 2008 |
|  | Funding: Hospital funds not reallocated to CMHS | Fakhoury & Priebe, 2002; PAHO, 2008 |
| 2. Knowledge / Science | Conceptual limitations and ambiguities | Bennett & Morris, 1983; Fakhoury & Priebe, 2002; Freedman & Moran, 1984; Mallik et al., 1998; McCubbin, 1994 |
|  | Evidence: Lack of evidence on DI processes | Shen & Snowden, 2014 |
|  | Lack of research and innovation on alternatives to institutionalization | Bennett & Morris, 1983 |
| 3. Power, interests, and influences | Irrelevance of Mental Health in the political/policy agenda | Mechanic & Rochefort, 1990; PAHO, 2008; Semke, 1999 |
|  | Market factors fostering re-institutionalization | Barton, 1983; Dorwart et al., 1991; Fakhoury & Priebe, 2002; Freedman & Moran, 1984; Rose, 1979 |
|  | Uncoordinated and fragmentary advocacy actions. | McCubbin, 1994; Mechanic & Rochefort, 1990; Rosenheck, 2000 |
|  | Vested interests: Pharmaceutical | McCubbin, 1994 |
| 4. Services and supports in the community | Centralized System | Kleiner & Drews, 1992 |
|  | Patients: Challenging behaviours | Allen et al., 2007 |
|  | Patients: Old Age | Barry et al., 2002 |
|  | Services: Hospital centric models and practices | Bennett & Morris, 1983; Kaffman et al., 1996 |
|  | Early discharge | Kleiner & Drews, 1992; Stelovich, 1979 |
|  | Services: Lack of services and support in the community | Fakhoury & Priebe, 2002; McCubbin, 1994; Oshima & Kuno, 2006; Weiss, 1990 |
|  | Housing: Inadequate, insufficient | Grabowski  et al., 2009; Mechanic & Rochefort, 1990; PAHO, 2008 |
|  | Dependence on disability benefits and/or pensions | Chopra & Herrman, 2011; Freedman & Moran, 1984; Manuel et al., 2012 |
|  | Patients: Disempowerment / Fatalism | Chopra & Herrman, 2011 |
|  | Insufficient Public Support | Manuel et al., 2012 |
|  | Patients: No money | Goering, 1984 |
|  | Clashing views on DI within the Workforce | Kleiner & Drews, 1992; PAHO, 2008 |
| 5. Workforce | Shortages in general | Fakhoury & Priebe, 2002; Schmidt, 2000; Shen & Snowden, 2014; WHO, 2014 |
|  | Shortages of specific professions | Ash et al., 2015 |
|  | Inadequate training | Barton, 1983; Mayston et al., 2016; PAHO, 2008; WHO, 2014 |
|  | Moral concerns and fears | Ash et al., 2015; Kleiner & Drews, 1992; PAHO, 2008 |
|  | Pessimism | Aggett & Goldberg, 2005; Cohen, 1983; Kleiner & Drews, 1992 |
|  | Practices of exclusion | Bryant et al., 2004; Chakraborty et al., 2011 |
|  | Stigma in workforce | Barton, 1983; Semke, 1999 |
|  | Vested interests: Workforce | Shen & Snowden, 2014; Swidler & Tauriello, 1995 |
| 6. Communities and the public | Communities are hostile towards users | Aggett & Goldberg, 2005; Bredenberg, 1983; Fakhoury & Priebe, 2002; O’Doherty et al., 2016; PAHO, 2008 |
|  | Communities are ill prepared to integrate users | Bredenberg, 1983; Fakhoury & Priebe, 2002 |
|  | Public acceptance of social control | Allen et al., 2007; Fakhoury & Priebe, 2002; Swidler & Tauriello, 1995 |
|  | Stigma & self-stigma | Aggett & Goldberg, 2005; Chan & Mak, 2014; Fakhoury & Priebe, 2002; Manuel et al., 2012; Mechanic & Rochefort, 1990; O’Doherty et al., 2016; PAHO, 2008 |
| 7. Family / Carers | Broken ties between families and services | Aggett & Goldberg, 2005 |
|  | Lack of support and/or unfair expectations towards families | Barton, 1983; Yip, 2006; Lavoie et al., 2012; Mechanic & Rochefort, 1990; Oshima & Kuno, 2006 |
|  | Scepticism and Opposition from families | McCubbin, 1994; Oshima & Kuno, 2006 |
