## Supplementary material for "Moving psychiatric deinstitutionalisation forward: A scoping review of barriers and facilitators": (12) Table 6 Facilitators

**Table 6: Facilitators to the process of psychiatric deinstitutionalization**

| **Category** | **Descriptive themes** | **References** |
| --- | --- | --- |
| Planning, Leadership and Funding/Economic aspects | Centralized governance of the process | PAHO, 2008 |
|  | Austerity and fiscal pressure | PAHO, 2008 |
|  | Disability insurance | Mechanic & Rochefort, 1990 |
|  | Economic incentives for DI | Mechanic & Rochefort, 1990 |
|  | Fiscal strain on state mental hospital | Mechanic & Rochefort, 1990; O’Doherty et al., 2016 |
|  | International policy networks and advocacy | PAHO, 2008 |
|  | Intersectoral alliances and coordination | PAHO, 2008 |
| Knowledge/Science | Available evidence about alternatives | Weiss, 1990 |
|  | Conceptual Clarity | Freedman & Moran, 1984; Kleiner & Drews, 1992; McCubbin, 1994 |
|  | Documented Experience | Shen & Snowden, 2014 |
|  | Evidence of human rights violations | PAHO, 2008 |
|  | Intellectual cross fertilization towards CBSs | Mechanic & Rochefort, 1990; PAHO, 2008 |
|  | Knowledge of effects of institutions on individual patients | Anderson et al., 1998; Bennett & Morris, 1983; Kleiner & Drews, 1992; Mechanic & Rochefort, 1990 |
|  | Psychopharmacological developments | Anderson et al., 1998; Bennett & Morris, 1983; Bredenberg, 1983; Freedman & Moran, 1984; Kleiner & Drews, 1992; Mechanic & Rochefort, 1990; Weiss, 1990 |
| Power, interests and influences | Human rights legislation | Anderson et al., 1998; PAHO, 2008 |
|  | Influence of civil rights movements | Mechanic & Rochefort, 1990; PAHO, 2008 |
|  | Legal limitations to commitment/coercion | Freedman & Moran, 1984; Mechanic & Rochefort, 1990; |
|  | Legal push towards community-based treatments | Freedman & Moran, 1984 |
|  | Legal standards for facility construction/operation | Anderson et al., 1998 |
|  | MH Legislation | Freedman & Moran, 1984; PAHO, 2008; Shen & Snowden, 2014 |
|  | Advocacy from professional organizations/groups | Weiss, 1990; WHO, 2014 |
|  | International policy pressure | Shen & Snowden, 2014 |
| Services and supports in the community | Service-user movements and demands | Anderson et al., 1998; Kleiner & Drews, 1992 |
|  | Comprehensive & structured network of CB services | Cohen, 1983; Conway et al., 1994; Evans et al., 2012; Lamb & Goertzel, 1977 |
|  | Continuity of care | Sytema et al., 1996 |
|  | Income for patients | Alakeson, 2010 |
|  | Individualization of care in the community | Kalisova et al., 2018 |
|  | Integration of mental health in PHC | Evans et al., 2012; John et al., 2014; Kraudy et al., 1987; PAHO, 2008 |
|  | Limit readmission by closing beds | PAHO, 2008 |
|  | Recovery-based services in a psych ICUs | Ash et al., 2015 |
|  | Scale up of outpatient services | Abas et al., 2003; Bennett & Morris, 1983 |
|  | Self-directed support: Autonomy in the use/selection of services | Alakeson, 2010 |
|  | Shared decision making and service user involvement | Chan & Mak, 2014 |
|  | Supporting PHC expertise to raise service-user confidence | Huang et al., 2017 |
|  | Social Help | Lamb & Goertzel; 1977 |
| Workforce | Anti-stigma practice | Huang et al., 2017; Matsea et al., 2019; Mayston et al., 2016 |
|  | PHC training | PAHO, 2008 |
|  | WF training | Wasylenki, 1995, Weiss, 1990 |
| Exogenous factors | Exogenous shocks (disasters, war) | Stelovich, 1979 |
|  | Re-democratization | Rizzardo et al., 1986 |
